## Supplementary material for "Covid-19 in end-stage renal disease patients with renal replacement therapies: a systematic review and meta-analysis": Funnel plot: estimated prevalence and case fatality rate with standard error of each included study in meta-analysis.

**S1 Fig. Funnel plot: estimated prevalence and case fatality rate with standard error of each included study in meta-analysis**

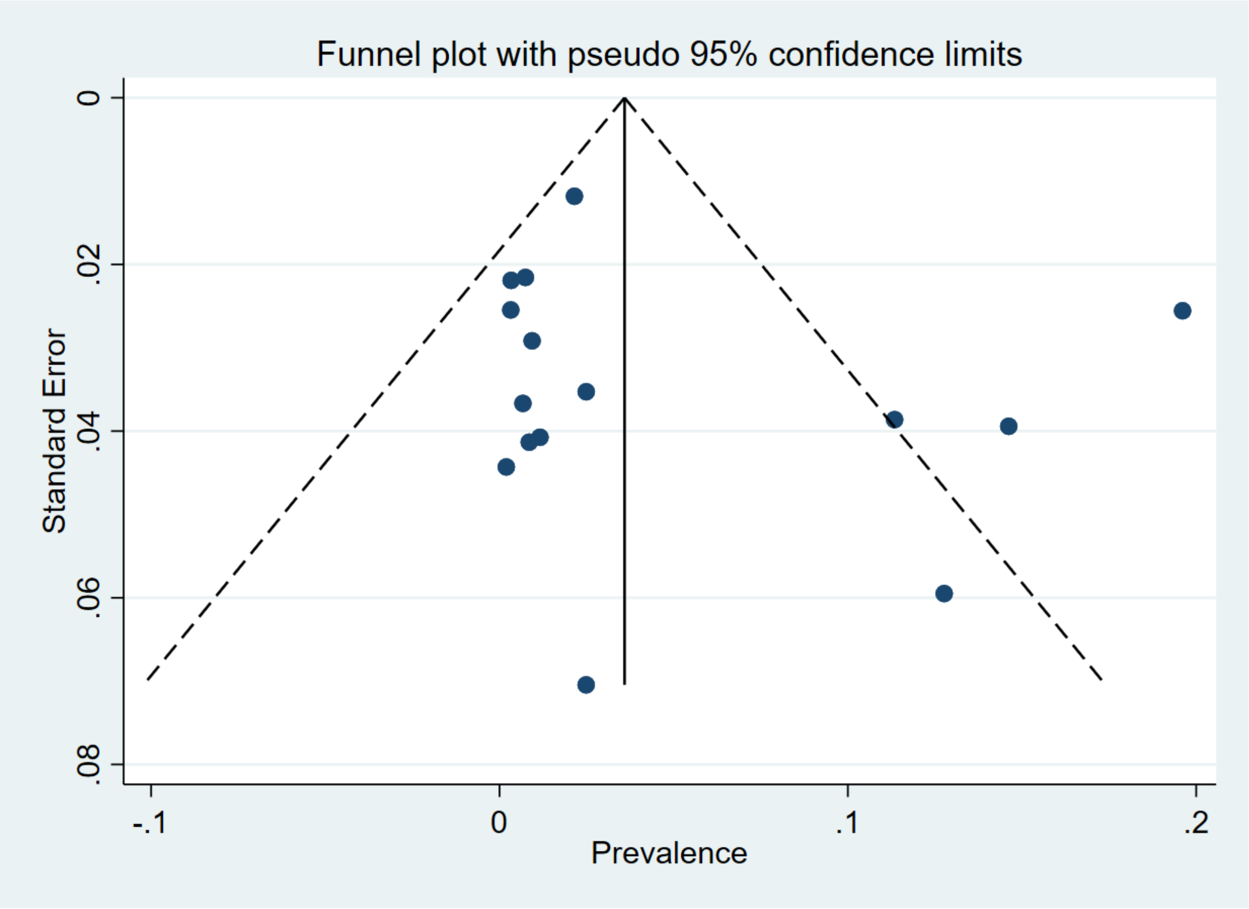


**S1A Fig. Funnel plot: estimated prevalence and standard error of each included study in meta-analysis.** Dark blue circles represent estimated prevalence for included studies. Vertical black line represents fixed effect estimated pooled prevalence. Sides of the triangle represent the expected 95% confidence intervals with inverted funnel shape.

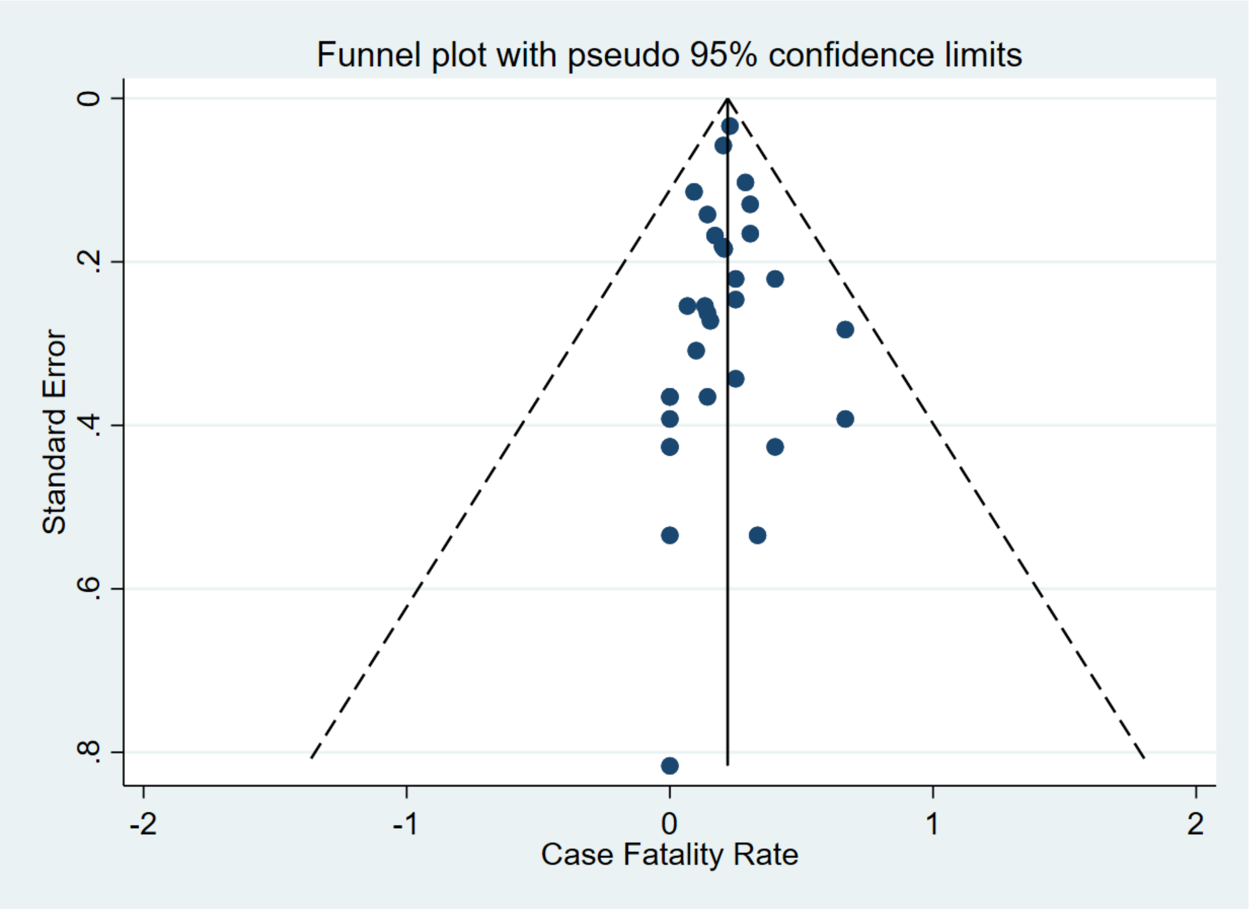


**S1B Fig. Funnel plot: estimated case fatality rate and standard error of each included study in meta-analysis.** Dark blue circles represent estimated case fatality rate for included studies. Vertical black line represents fixed effect estimated pooled case fatality rate. Sides of the triangle represent the expected 95% confidence intervals with inverted funnel.
