## Supplementary material for "Covid-19 in end-stage renal disease patients with renal replacement therapies: a systematic review and meta-analysis": Risk of bias assessment

**S1 Table. Risk of bias assessment**

**S1A Table. Risk of bias assessment for included studies with prevalence outcomes.**

| **Author** | **Sample Representative**  **ness** | **Sampling Frame** | **Sampling Techniques** | **Response Rate** | **Data Collection Method** | **Case Definition** | **Measurement Tools** | **Mode of Measurement** | **Study Period** | **Data Calculation** | **Overall** |
| --- | --- | --- | --- | --- | --- | --- | --- | --- | --- | --- | --- |
| Akdur^25^ | High | Low | Low | Low | Low | Low | Low | Low | Low | Low | Mild |
| Alberici a^19^ | High | Low | Low | Low | Low | Low | Low | Low | Low | Low | Mild |
| Arslan^26^ | High | High | High | High | Low | Low | Low | Low | Low | Low | Moderate |
| Banerjee^27^ | High | Low | Low | Low | Low | Low | Low | Low | Low | Low | Mild |
| Cho^22^ | High | Low | Low | Low | Low | Low | Low | Low | Low | Low | Mild |
| Corbett^28^ | High | Low | Low | Low | Low | Low | Low | Low | Low | Low | Mild |
| Crespo^23^ | High | Low | Low | Low | Low | Low | Low | Low | Low | Low | Mild |
| Goicoechea^24^ | High | Low | Low | Low | Low | Low | Low | Low | Low | Low | Mild |
| Hoek^21^ | High | Low | Low | Low | Low | Low | Low | Low | Low | Low | Mild |
| Maritati^20^ | High | Low | Low | Low | Low | Low | Low | Low | Low | Low | Mild |
| Roper^29^ | High | High | High | High | Low | Low | Low | Low | Low | Low | Moderate |
| Wang^15^ | High | Low | Low | Low | Low | Low | Low | Low | Low | Low | Mild |
| Xiong^16^ | High | Low | Low | Low | Low | Low | Low | Low | Low | Low | Mild |
| Xu^17^ | High | Low | Low | Low | Low | Low | Low | Low | Low | Low | Mild |
| Zhang^18^ | High | High | High | High | Low | Low | Low | Low | Low | Low | Moderate |

**S1B Table. Risk of bias assessment for included studies with case fatality rate outcomes.**

| **Author** | **Inclusion of Consecutive Cases** | **Multicenter** | **More than 80% Follow-up** | **Multivariable Analysis** |
| --- | --- | --- | --- | --- |
| Abrishami^33^ | Adequate | Inadequate | Adequate | Inadequate |
| Akdur^25^ | Adequate | Inadequate | Adequate | Inadequate |
| Alberici a^19^ | Adequate | Adequate | Adequate | Adequate |
| Alberici b^34^ | Adequate | Inadequate | Adequate | Inadequate |
| Arslan^26^ | Inadequate | Adequate | Adequate | Inadequate |
| Banerjee^27^ | Adequate | Inadequate | Adequate | Inadequate |
| Bösch^32^ | Adequate | Inadequate | Adequate | Inadequate |
| Chen^43^ | Adequate | Inadequate | Adequate | Inadequate |
| Corbett^28^ | Adequate | Inadequate | Adequate | Adequate |
| Columbia U^44^ | Adequate | Inadequate | Adequate | Inadequate |
| Crespo^23^ | Adequate | Inadequate | Adequate | Inadequate |
| Fernández-Ruiz^38^ | Adequate | Inadequate | Adequate | Inadequate |
| Fung^45^ | Adequate | Inadequate | Adequate | Inadequate |
| Goicoechea^24^ | Adequate | Inadequate | Adequate | Inadequate |
| Hoek^21^ | Adequate | Adequate | Adequate | Inadequate |
| Jung^37^ | Adequate | Adequate | Adequate | Inadequate |
| Kolonko^36^ | Adequate | Inadequate | Adequate | Inadequate |
| Maritati^20^ | Adequate | Inadequate | Adequate | Inadequate |
| Mehta^46^ | Adequate | Inadequate | Adequate | Inadequate |
| Melgosa^39^ | Adequate | Inadequate | Adequate | Inadequate |
| Mella^35^ | Adequate | Inadequate | Adequate | Inadequate |
| Rodriguez-Cubillo^40^ | Adequate | Inadequate | Adequate | Inadequate |
| Roper^29^ | Adequate | Inadequate | Adequate | Inadequate |
| Sánchez-Álvarez^41^ | Adequate | Adequate | Adequate | Adequate |
| Tschopp^42^ | Adequate | Inadequate | Adequate | Inadequate |
| Valeri^7^ | Adequate | Inadequate | Adequate | Inadequate |
| Wang^15^ | Adequate | Inadequate | Adequate | Inadequate |
| Wu^30^ | Adequate | Inadequate | Adequate | Inadequate |
| Yi^47^ | Adequate | Inadequate | Adequate | Inadequate |
| Zhang^18^ | Adequate | Inadequate | Adequate | Inadequate |
| Zhu^31^ | Adequate | Inadequate | Adequate | Inadequate |
