## Supplementary material for "Covid-19 in end-stage renal disease patients with renal replacement therapies: a systematic review and meta-analysis": Forest plots with subgroup analysis: mechanical ventilation rate

### S2 Fig. Forest plots with subgroup analysis: mechanical ventilation rate

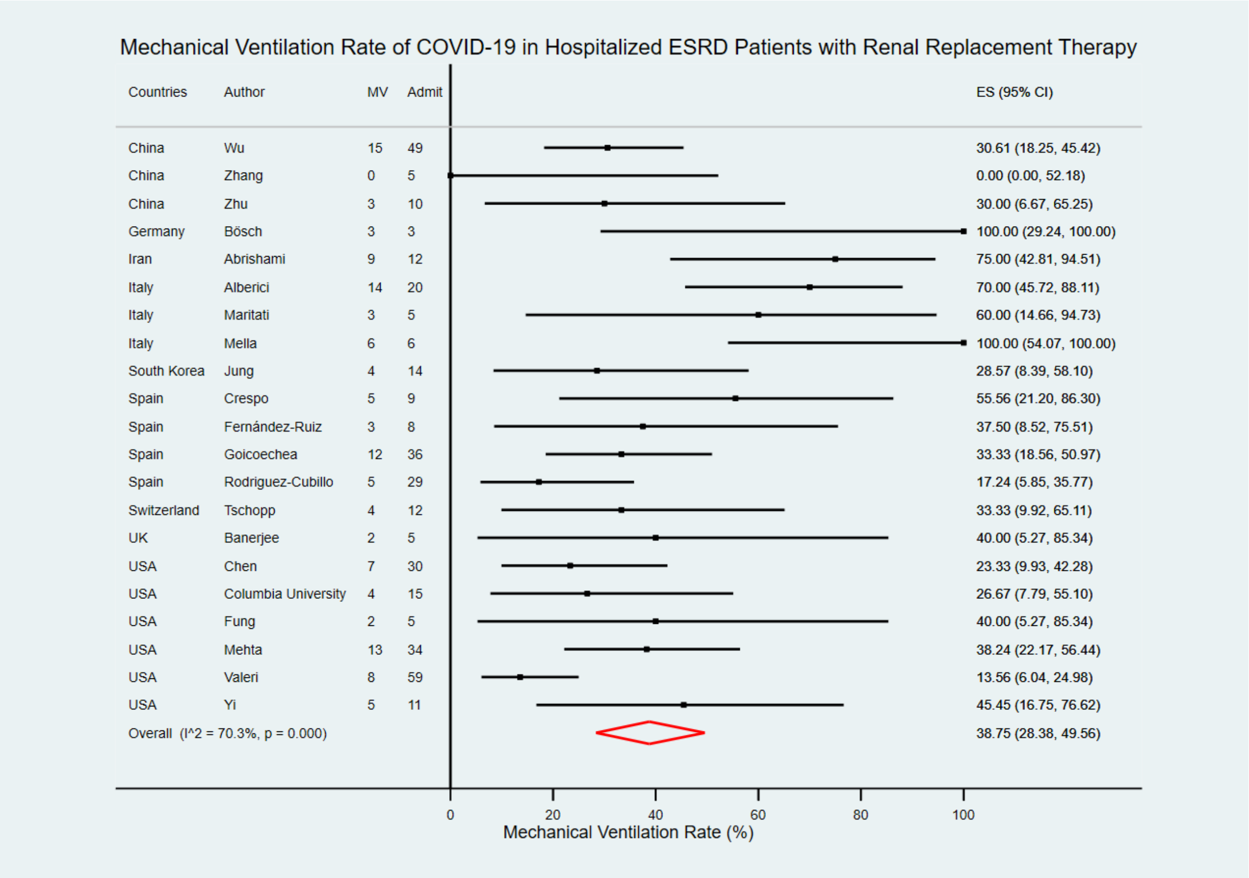


**S2A Fig. Forest plot: mechanical ventilation rate of COVID-19 in hospitalized ESRD patients with RRT.** The figure summarizes the number of hospitalized COVID-19 cases with need of mechanical ventilation in ESRD patients with RRT and the number of hospitalized COVID-19 cases in ESRD patients with RRT in 21 eligible studies. The forest plot represents the estimated mechanical ventilation rate of COVID-19 in hospitalized ESRD patients with RRT for each study (black boxes), with 95% confidence intervals (95% CI; horizontal black lines). The overall estimated pooled mechanical ventilation rate (red diamond) was 38.75% (95% CI = 28.38, 49.56%). The meta-analysis used a random-effects model with the exact method for confidence interval estimation. ES, effect size. $I^2$, test for heterogeneity.

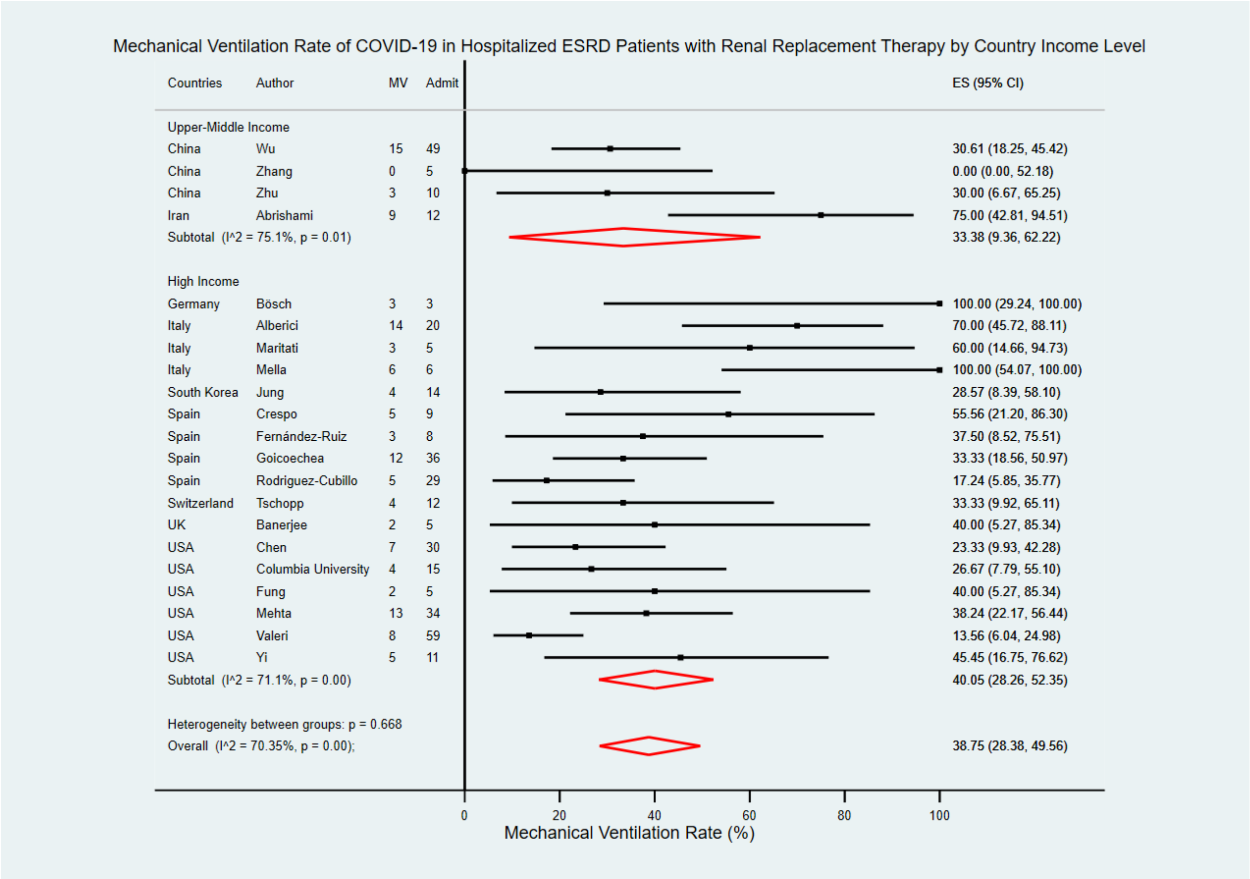


**S2B Fig. Forest plot: mechanical ventilation rate of COVID-19 in hospitalized ESRD patients with RRT by country income level.** The figure summarizes the number of hospitalized COVID-19 cases with need of mechanical ventilation in ESRD patients with RRT and the number of hospitalized COVID-19 cases in ESRD patients with RRT in 21 eligible studies with subgroup analysis by the World Bank country income level. The forest plot represents the estimated mechanical ventilation rate of COVID-19 in hospitalized ESRD patients with RRT for each study (black boxes), with 95% confidence intervals (95% CI; horizontal black lines). The estimated pooled mechanical ventilation rate for each subgroup was presented with a red diamond. The overall estimated pooled mechanical ventilation rate (last red diamond) was 38.75% (95% CI = 28.38, 49.56%). The meta-analysis used a random-effects model with the exact method for confidence interval estimation. ES, effect size. $I^2$, test for heterogeneity.

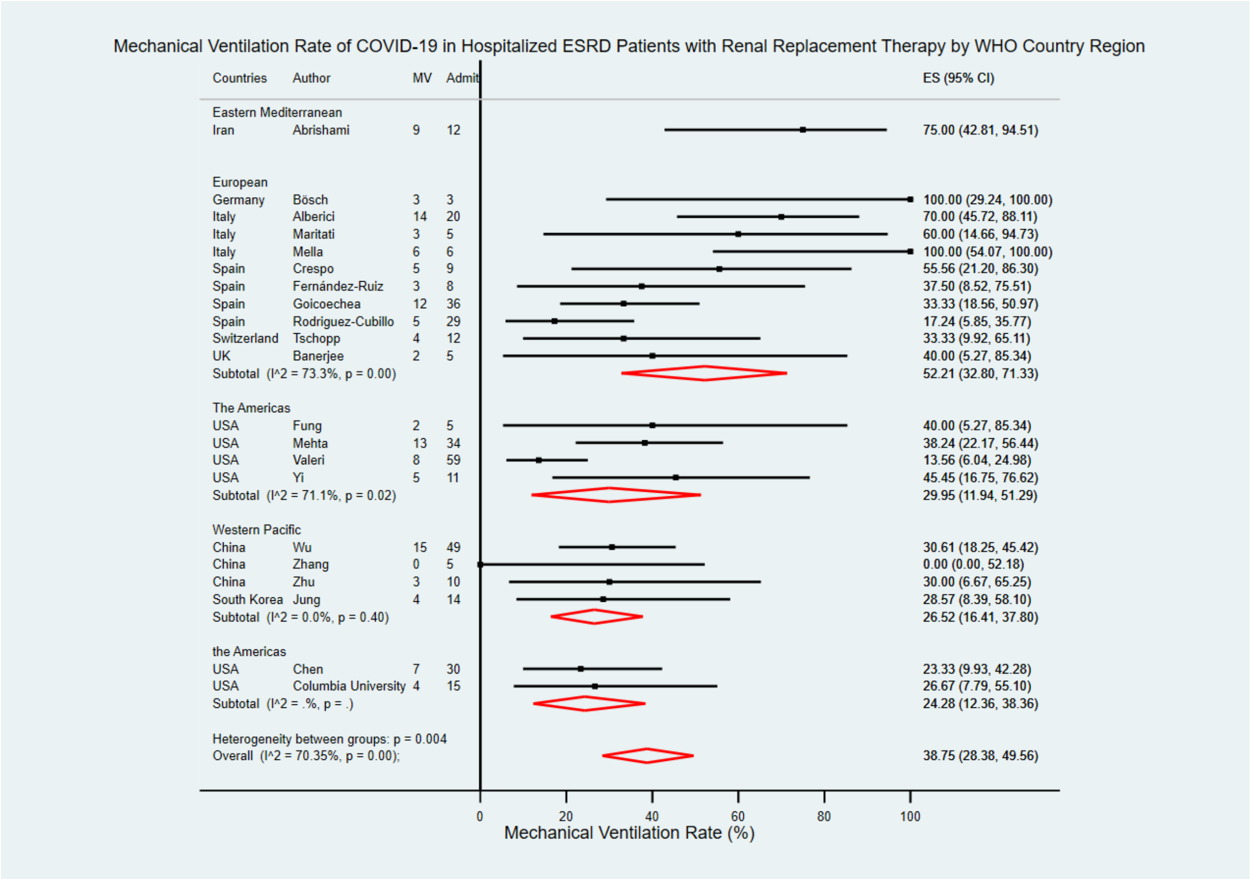


**S2C Fig. Forest plot: mechanical ventilation rate of COVID-19 in hospitalized ESRD patients with RRT by WHO country region.** The figure summarizes the number of hospitalized COVID-19 cases with need of mechanical ventilation in ESRD patients with RRT and the number of hospitalized COVID-19 cases in ESRD patients with RRT in 21 eligible studies with subgroup analysis by WHO country regions. The forest plot represents the estimated mechanical ventilation rate of COVID-19 in hospitalized ESRD patients with RRT for each study (black boxes), with 95% confidence intervals (95% CI; horizontal black lines). The estimated pooled mechanical ventilation rate for each subgroup was presented with a red diamond. The overall estimated pooled mechanical ventilation rate (last red diamond) was 38.75% (95% CI = 28.38, 49.56%). The meta-analysis used a random-effects model with the exact method for confidence interval estimation. ES, effect size. $I^2$, test for heterogeneity.

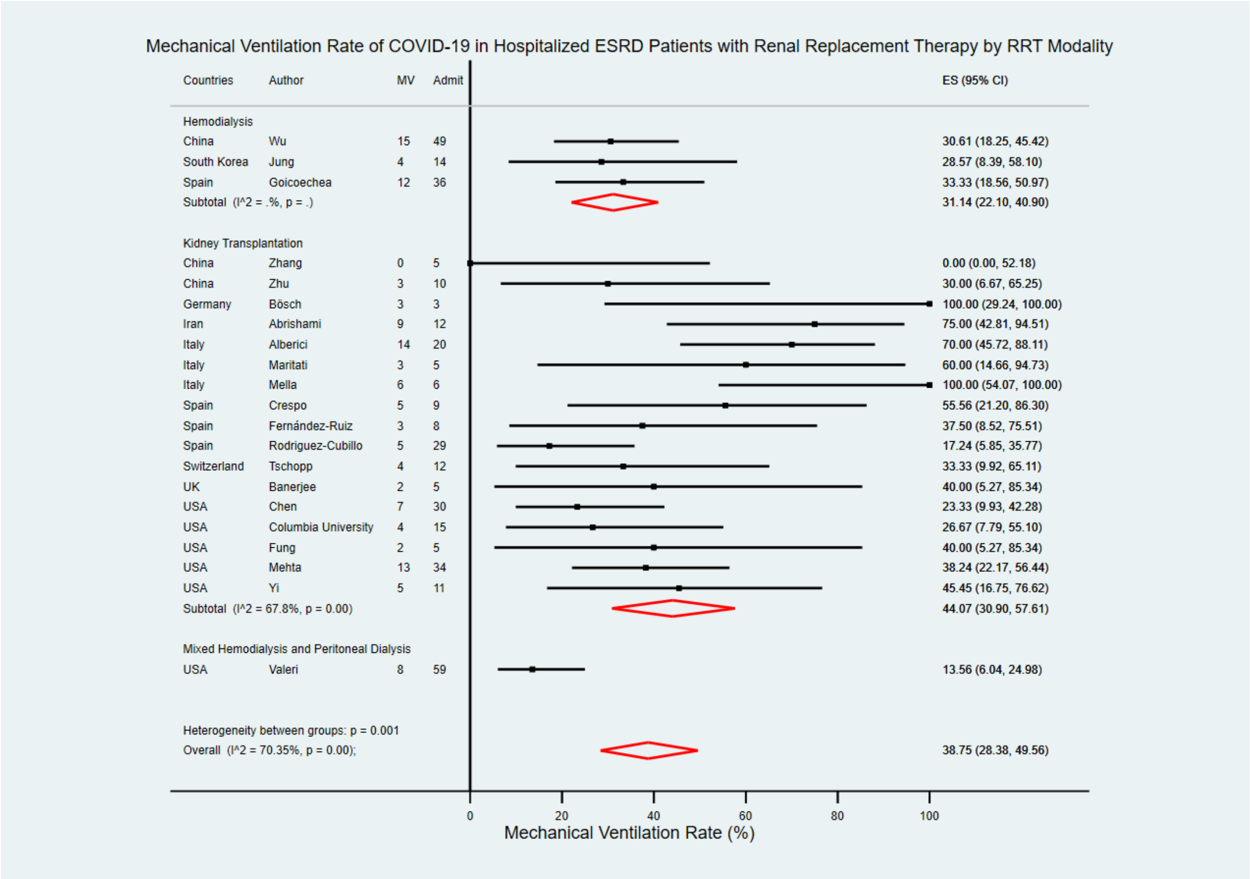


**S2D Fig. Forest plot: mechanical ventilation rate of COVID-19 in hospitalized ESRD patients with RRT by RRT modality.** The figure summarizes the number of hospitalized COVID-19 cases with need of mechanical ventilation in ESRD patients with RRT and the number of hospitalized COVID-19 cases in ESRD patients with RRT in 21 eligible studies with subgroup analysis by types of RRT modality. The forest plot represents the estimated mechanical ventilation rate of COVID-19 in hospitalized ESRD patients with RRT for each study (black boxes), with 95% confidence intervals (95% CI; horizontal black lines). The estimated pooled mechanical ventilation rate for each subgroup was presented with a red diamond. The overall estimated pooled mechanical ventilation rate (last red diamond) was 38.75% (95% CI = 28.38, 49.56%). The meta-analysis used a random-effects model with the exact method for confidence interval estimation. ES, effect size. $I^2$, test for heterogeneity.
