## Supplementary material for "Covid-19 in end-stage renal disease patients with renal replacement therapies: a systematic review and meta-analysis": Full search strategy

**S2 Table. Full search strategy**

**S2A Table. Full search strategy for PubMed.**

| Set # | PubMed | Results |
| --- | --- | --- |
| 1  COVID-19 | "coronavirus"[MeSH Terms] OR "coronavirus infections"[MeSH Terms] OR "betacoronavirus"[MeSH Terms] OR (COVID[tiab] AND 19[tiab]) OR "COVID-19"[tiab] OR "COVID 19"[tiab] OR COVID19[tiab] OR (corona*[tiab] AND (virus*[tiab] OR viral*[tiab] OR virinae*[tiab])) OR coronavirus*[tiab] OR coronaviral*[tiab] OR coronavirinae*[tiab] OR "novel coronavirus"[tiab] OR "new coronavirus"[tiab] OR (2019[tiab] AND nCoV[tiab]) OR "2019-nCoV"[tiab] OR "2019 nCoV"[tiab] OR "Wuhan coronavirus"[tiab] OR "SARS-CoV-2"[tiab] OR "SARS CoV 2"[tiab] OR "SARSCoV2"[tiab] | **48593** |
| 2  Renal Replacement  Therapy | "renal replacement therapy"[MeSH Terms] OR ((renal[tiab] OR kidney*[tiab]) AND replace*[tiab] AND therap*[tiab]) OR "renal replacement therapy"[tiab] OR "renal replacement therapies"[tiab] OR "kidney replacement therapy"[tiab] OR "kidney replacement therapies"[tiab] OR RRT[tiab] | **220228** |
| 3 Dialysis | "renal dialysis"[MeSH Terms] OR dialysis[tiab] OR dialyses[tiab] | **161812** |
| 4  Peritoneal Dialysis | "peritoneal dialysis"[MeSH Terms] OR (peritoneal[tiab] AND (dialysis[tiab] OR dialyses[tiab])) OR "peritoneal dialysis"[tiab] OR "peritoneal dialyses"[tiab] OR pd[tiab] OR capd[tiab] OR ccpd[tiab] OR apd[tiab] | **164014** |
| 5  Hemodialysis | hemodialysis[tiab] OR hemodialyses[tiab] OR haemodialysis[tiab] OR haemodialyses[tiab] OR hd[tiab] OR "hemofiltration"[MeSH Terms] OR hemofiltration[tiab] OR haemofiltration[tiab] OR "hemodiafiltration"[MeSH Terms] OR hemodiafiltration[tiab] OR haemodiafiltration[tiab] | **113882** |
| 6  Kidney Transplant | "kidney transplantation"[MeSH Terms] OR ((renal[tiab] OR kidney[tiab]) AND (transplant*[tiab] OR graft*[tiab])) OR "kidney transplant"[tiab] OR "kidney transplants"[tiab] OR "kidney transplantation"[tiab] OR "kidney transplantations"[tiab] OR "renal transplant"[tiab] OR "renal transplants"[tiab] OR "renal transplantation"[tiab] OR "renal transplantations"[tiab] | **141917** |
| 7  ESRD | "kidney failure, chronic"[MeSH Terms] OR ((chronic[tiab] OR (end[tiab] AND stage*[tiab]) OR endstage*[tiab]) AND (kidney[tiab] OR renal[tiab]) AND (fail*[tiab] OR disease*[tiab])) OR "chronic kidney failure"[tiab] OR "chronic kidney failures"[tiab] OR "chronic renal failure"[tiab] OR "chronic renal failures"[tiab] OR "chronic kidney disease"[tiab] OR "chronic kidney diseases"[tiab] OR "end-stage kidney disease"[tiab] OR "end-stage kidney diseases"[tiab] OR "end stage kidney disease"[tiab] OR "end stage kidney diseases"[tiab] OR "endstage kidney disease"[tiab] OR "endstage kidney diseases"[tiab] OR "end-stage renal disease"[tiab] OR "end-stage renal diseases"[tiab] OR "end stage renal disease"[tiab] OR "end stage renal diseases"[tiab] OR "endstage renal disease"[tiab] OR "endstage renal diseases"[tiab] OR CRF[tiab] OR CRD[tiab] OR CKF[tiab] OR CKD[tiab] OR ESRF[tiab] OR ESRD[tiab] OR ESKF[tiab] OR ESKD[tiab] OR ((kidney[tiab] OR renal[tiab]) AND insufficien*[tiab]) | **221733** |
| 8 | #2 OR #3 OR #4 OR #5 OR #6 OR #7 | **592526** |
| 9 | #1 AND #8 | **735** |
| 10 | animals[MeSH Terms] NOT humans[MeSH Terms] | **4712175** |
| 11 | #9 NOT #10 | **699** |
| 12 | english[lang] | **26448828** |
| 13 | #11 AND #12 | **665** |

**S2B Table. Full search strategy for Embase.**

| Set # | Embase | Results |
| --- | --- | --- |
| 1  COVID-19 | 'coronavirinae'/exp OR 'coronavirus disease 2019'/exp OR 'betacoronavirus'/exp OR ('COVID':ti,ab AND '19':ti,ab) OR 'COVID-19':ti,ab OR 'COVID 19':ti,ab OR 'COVID19':ti,ab OR ('corona*':ti,ab AND ('virus*':ti,ab OR 'viral*':ti,ab OR 'virinae*':ti,ab)) OR 'coronavirus*':ti,ab OR 'coronaviral*':ti,ab OR 'coronavirinae*':ti,ab OR 'novel coronavirus':ti,ab OR 'new coronavirus':ti,ab OR ('2019':ti,ab AND 'nCoV':ti,ab) OR '2019-nCoV':ti,ab OR '2019 nCoV':ti,ab OR 'Wuhan coronavirus':ti,ab OR 'severe acute respiratory syndrome coronavirus 2'/exp OR 'SARS-CoV-2':ti,ab OR 'SARS CoV 2':ti,ab OR 'SARSCoV2':ti,ab | **43063** |
| 2  Renal Replacement  Therapy | 'renal replacement therapy'/exp OR (('renal':ti,ab OR 'kidney*':ti,ab) AND 'replace*':ti,ab AND 'therap*':ti,ab) OR 'renal replacement therapy':ti,ab OR 'renal replacement therapies':ti,ab OR 'kidney replacement therapy':ti,ab OR 'kidney replacement therapies':ti,ab OR 'RRT':ti,ab | **204903** |
| 3 Dialysis | 'dialysis':ti,ab OR 'dialyses':ti,ab | **149890** |
| 4  Peritoneal Dialysis | 'peritoneal dialysis'/exp OR ('peritoneal':ti,ab AND ('dialysis':ti,ab OR 'dialyses':ti,ab)) OR 'peritoneal dialysis':ti,ab OR 'peritoneal dialyses':ti,ab OR 'pd':ti,ab OR 'capd':ti,ab OR 'ccpd':ti,ab OR 'apd':ti,ab | **261886** |
| 5  Hemodialysis | 'hemodialysis'/exp OR 'hemodialysis':ti,ab OR 'hemodialyses':ti,ab OR 'haemodialysis':ti,ab OR 'haemodialyses':ti,ab OR 'hd':ti,ab OR 'hemofiltration'/exp OR 'hemofiltration':ti,ab OR 'haemofiltration':ti,ab OR 'hemodiafiltration'/exp OR 'hemodiafiltration':ti,ab OR 'haemodiafiltration':ti,ab | **195579** |
| 6  Kidney Transplant | 'kidney transplantation'/exp OR (('renal':ti,ab OR 'kidney':ti,ab) AND ('transplant*':ti,ab OR 'graft*':ti,ab)) OR 'kidney transplant':ti,ab OR 'kidney transplants':ti,ab OR 'kidney transplantation':ti,ab OR 'kidney transplantations':ti,ab OR 'renal transplant':ti,ab OR 'renal transplants':ti,ab OR 'renal transplantation':ti,ab OR 'renal transplantations':ti,ab | **220864** |
| 7  ESRD | 'chronic kidney failure'/exp OR 'end stage renal disease'/exp OR (('chronic':ti,ab OR ('end':ti,ab AND 'stage*':ti,ab) OR 'endstage*':ti,ab) AND ('kidney':ti,ab OR 'renal':ti,ab) AND ('fail*':ti,ab OR 'disease*':ti,ab)) OR 'chronic kidney failure':ti,ab OR 'chronic kidney failures':ti,ab OR 'chronic renal failure':ti,ab OR 'chronic renal failures':ti,ab OR 'chronic kidney disease':ti,ab OR 'chronic kidney diseases':ti,ab OR 'end-stage kidney disease':ti,ab OR 'end-stage kidney diseases':ti,ab OR 'end stage kidney disease':ti,ab OR 'end stage kidney diseases':ti,ab OR 'endstage kidney disease':ti,ab OR 'endstage kidney diseases':ti,ab OR 'end-stage renal disease':ti,ab OR 'end-stage renal diseases':ti,ab OR 'end stage renal disease':ti,ab OR 'end stage renal diseases':ti,ab OR 'endstage renal disease':ti,ab OR 'endstage renal diseases':ti,ab OR 'CRF':ti,ab OR 'CRD':ti,ab OR 'CKF':ti,ab OR 'CKD':ti,ab OR 'ESRF':ti,ab OR 'ESRD':ti,ab OR 'ESKF':ti,ab OR 'ESKD':ti,ab OR (('kidney':ti,ab OR 'renal':ti,ab) AND 'insufficien*':ti,ab) | **322949** |
| 8 | #2 OR #3 OR #4 OR #5 OR #6 OR #7 | **901345** |
| 9 | #1 AND #8 | **892** |
| 10 | [animals]/lim NOT [humans]/lim | **58000732** |
| 11 | #9 NOT #10 | **844** |
| 12 | english:la | **31385730** |
| 13 | #11 AND #12 | **815** |

**S2C Table. Full search strategy for Scopus.**

| Set # | Scopus | Results |
| --- | --- | --- |
| 1  COVID-19 | TITLE-ABS-KEY((COVID AND 19) OR "COVID-19" OR "COVID 19" OR COVID19 OR (corona* AND (virus* OR viral* OR virinae*)) OR coronavirus* OR coronaviral* OR coronavirinae* OR "novel coronavirus" OR "new coronavirus" OR (2019 AND nCoV) OR "2019-nCoV" OR "2019 nCoV" OR "Wuhan coronavirus" OR "SARS-CoV-2" OR "SARS CoV 2" OR "SARSCoV2") | **50337** |
| 2  Renal Replacement  Therapy | TITLE-ABS-KEY(((renal OR kidney*) AND replace* AND therap*) OR "renal replacement therapy" OR "renal replacement therapies" OR "kidney replacement therapy" OR "kidney replacement therapies" OR RRT) | **59250** |
| 3 Dialysis | TITLE-ABS-KEY(dialysis OR dialyses) | **206186** |
| 4  Peritoneal Dialysis | TITLE-ABS-KEY((peritoneal AND (dialysis OR dialyses)) OR "peritoneal dialysis" OR "peritoneal dialyses" OR pd OR capd OR ccpd OR apd) | **351310** |
| 5  Hemodialysis | TITLE-ABS-KEY(hemodialysis OR hemodialyses OR haemodialysis OR haemodialyses OR hd OR hemofiltration OR haemofiltration OR hemodiafiltration OR haemodiafiltration) | **212779** |
| 6  Kidney Transplant | TITLE-ABS-KEY(((renal OR kidney) AND (transplant* OR graft*)) OR "kidney transplant" OR "kidney transplants" OR "kidney transplantation" OR "kidney transplantations" OR "renal transplant" OR "renal transplants" OR "renal transplantation" OR "renal transplantations") | **207136** |
| 7  ESRD | TITLE-ABS-KEY(((chronic OR (end AND stage*) OR endstage*) AND (kidney OR renal) AND (fail* OR disease*)) OR "chronic kidney failure" OR "chronic kidney failures" OR "chronic renal failure" OR "chronic renal failures" OR "chronic kidney disease" OR "chronic kidney diseases" OR "end-stage kidney disease" OR "end-stage kidney diseases" OR "end stage kidney disease" OR "end stage kidney diseases" OR "endstage kidney disease" OR "endstage kidney diseases" OR "end-stage renal disease" OR "end-stage renal diseases" OR "end stage renal disease" OR "end stage renal diseases" OR "endstage renal disease" OR "endstage renal diseases" OR CRF OR CRD OR CKF OR CKD OR ESRF OR ESRD OR ESKF OR ESKD OR ((kidney OR renal) AND insufficien*)) | **335740** |
| 8 | #2 OR #3 OR #4 OR #5 OR #6 OR #7 | **998618** |
| 9 | #1 AND #8 | **1381** |
| 10 | ALL(animals AND NOT humans) | **3962658** |
| 11 | #9 AND NOT #10 | **1351** |
| 12 | LANGUAGE(english) | **68149064** |
| 13 | #11 AND #12 | **1297** |

**S2D Table. Full search strategy for Web of Science.**

| Set # | Web of Science | Results |
| --- | --- | --- |
| 1  COVID-19 | TS=((COVID AND 19) OR "COVID-19" OR "COVID 19" OR COVID19 OR (corona* AND (virus* OR viral* OR virinae*)) OR coronavirus* OR coronaviral* OR coronavirinae* OR "novel coronavirus" OR "new coronavirus" OR (2019 AND nCoV) OR "2019-nCoV" OR "2019 nCoV" OR "Wuhan coronavirus" OR "SARS-CoV-2" OR "SARS CoV 2" OR "SARSCoV2") | **28674** |
| 2  Renal Replacement  Therapy | TS=(((renal OR kidney*) AND replace* AND therap*) OR "renal replacement therapy" OR "renal replacement therapies" OR "kidney replacement therapy" OR "kidney replacement therapies" OR RRT) | **22322** |
| 3 Dialysis | TS=(dialysis OR dialyses) | **104884** |
| 4  Peritoneal Dialysis | TS=((peritoneal AND (dialysis OR dialyses)) OR "peritoneal dialysis" OR "peritoneal dialyses" OR pd OR capd OR ccpd OR apd) | **263489** |
| 5  Hemodialysis | TS=(hemodialysis OR hemodialyses OR haemodialysis OR haemodialyses OR hd OR hemofiltration OR haemofiltration OR hemodiafiltration OR haemodiafiltration) | **129970** |
| 6  Kidney Transplant | TS=(((renal OR kidney) AND (transplant* OR graft*)) OR "kidney transplant" OR "kidney transplants" OR "kidney transplantation" OR "kidney transplantations" OR "renal transplant" OR "renal transplants" OR "renal transplantation" OR "renal transplantations") | **142335** |
| 7  ESRD | TS=(((chronic OR (end AND stage*) OR endstage*) AND (kidney OR renal) AND (fail* OR disease*)) OR "chronic kidney failure" OR "chronic kidney failures" OR "chronic renal failure" OR "chronic renal failures" OR "chronic kidney disease" OR "chronic kidney diseases" OR "end-stage kidney disease" OR "end-stage kidney diseases" OR "end stage kidney disease" OR "end stage kidney diseases" OR "endstage kidney disease" OR "endstage kidney diseases" OR "end-stage renal disease" OR "end-stage renal diseases" OR "end stage renal disease" OR "end stage renal diseases" OR "endstage renal disease" OR "endstage renal diseases" OR CRF OR CRD OR CKF OR CKD OR ESRF OR ESRD OR ESKF OR ESKD OR ((kidney OR renal) AND insufficien*)) | **186498** |
| 8 | #2 OR #3 OR #4 OR #5 OR #6 OR #7 | **675668** |
| 9 | #1 AND #8 | **508** |
| 10 | ALL=(animal NOT human) | **1086780** |
| 11 | #9 NOT #10 | **487** |
| 12 | #11 restrict language to English | **471** |

**S2E Table. Full search strategy for CENTRAL.**

| Set # | CENTRAL | Results |
| --- | --- | --- |
| 1  COVID-19 | [mh coronavirus] OR [mh "coronavirus infections"] OR [mh betacoronavirus] OR (COVID:ti,ab,kw AND 19:ti,ab,kw) OR "COVID-19":ti,ab,kw OR "COVID 19":ti,ab,kw OR COVID19:ti,ab,kw OR (corona*:ti,ab,kw AND (virus*:ti,ab,kw OR viral*:ti,ab,kw OR virinae*:ti,ab,kw)) OR coronavirus*:ti,ab,kw OR coronaviral*:ti,ab,kw OR coronavirinae*:ti,ab,kw OR "novel coronavirus":ti,ab,kw OR "new coronavirus":ti,ab,kw OR (2019:ti,ab,kw AND nCoV:ti,ab,kw) OR "2019-nCoV":ti,ab,kw OR "2019 nCoV":ti,ab,kw OR "Wuhan coronavirus":ti,ab,kw OR "SARS-CoV-2":ti,ab,kw OR "SARS CoV 2":ti,ab,kw OR "SARSCoV2":ti,ab,kw | **917** |
| 2  Renal Replacement  Therapy | [mh "renal replacement therapy"] OR ((renal:ti,ab,kw OR kidney*:ti,ab,kw) AND replace*:ti,ab,kw AND therap*:ti,ab,kw) OR "renal replacement therapy":ti,ab,kw OR "renal replacement therapies":ti,ab,kw OR "kidney replacement therapy":ti,ab,kw OR "kidney replacement therapies":ti,ab,kw OR RRT:ti,ab,kw | **11237** |
| 3 Dialysis | [mh "renal dialysis"] OR dialysis:ti,ab,kw OR dialyses:ti,ab,kw | **13351** |
| 4  Peritoneal Dialysis | [mh "peritoneal dialysis"] OR (peritoneal:ti,ab,kw AND (dialysis:ti,ab,kw OR dialyses:ti,ab,kw)) OR "peritoneal dialysis":ti,ab,kw OR "peritoneal dialyses":ti,ab,kw OR pd:ti,ab,kw OR capd:ti,ab,kw OR ccpd:ti,ab,kw OR apd:ti,ab,kw | **35920** |
| 5  Hemodialysis | hemodialysis:ti,ab,kw OR hemodialyses:ti,ab,kw OR haemodialysis:ti,ab,kw OR haemodialyses:ti,ab,kw OR hd:ti,ab,kw OR [mh hemofiltration] OR hemofiltration:ti,ab,kw OR haemofiltration:ti,ab,kw OR [mh hemodiafiltration] OR hemodiafiltration:ti,ab,kw OR haemodiafiltration:ti,ab,kw | **15278** |
| 6  Kidney Transplant | [mh "kidney transplantation"] OR ((renal:ti,ab,kw OR kidney:ti,ab,kw) AND (transplant*:ti,ab,kw OR graft*:ti,ab,kw)) OR "kidney transplant":ti,ab,kw OR "kidney transplants":ti,ab,kw OR "kidney transplantation":ti,ab,kw OR "kidney transplantations":ti,ab,kw OR "renal transplant":ti,ab,kw OR "renal transplants":ti,ab,kw OR "renal transplantation":ti,ab,kw OR "renal transplantations":ti,ab,kw | **14291** |
| 7  ESRD | [mh "kidney failure, chronic"] OR ((chronic:ti,ab,kw OR (end:ti,ab,kw AND stage*:ti,ab,kw) OR endstage*:ti,ab,kw) AND (kidney:ti,ab,kw OR renal:ti,ab,kw) AND (fail*:ti,ab,kw OR disease*:ti,ab,kw)) OR "chronic kidney failure":ti,ab,kw OR "chronic kidney failures":ti,ab,kw OR "chronic renal failure":ti,ab,kw OR "chronic renal failures":ti,ab,kw OR "chronic kidney disease":ti,ab,kw OR "chronic kidney diseases":ti,ab,kw OR "end-stage kidney disease":ti,ab,kw OR "end-stage kidney diseases":ti,ab,kw OR "end stage kidney disease":ti,ab,kw OR "end stage kidney diseases":ti,ab,kw OR "endstage kidney disease":ti,ab,kw OR "endstage kidney diseases":ti,ab,kw OR "end-stage renal disease":ti,ab,kw OR "end-stage renal diseases":ti,ab,kw OR "end stage renal disease":ti,ab,kw OR "end stage renal diseases":ti,ab,kw OR "endstage renal disease":ti,ab,kw OR "endstage renal diseases":ti,ab,kw OR CRF:ti,ab,kw OR CRD:ti,ab,kw OR CKF:ti,ab,kw OR CKD:ti,ab,kw OR ESRF:ti,ab,kw OR ESRD:ti,ab,kw OR ESKF:ti,ab,kw OR ESKD:ti,ab,kw OR ((kidney:ti,ab,kw OR renal:ti,ab,kw) AND insufficien*:ti,ab,kw) | **24966** |
| 8 | #2 OR #3 OR #4 OR #5 OR #6 OR #7 | **81231** |
| 9 | #1 AND #8 | **74** |
| 10 | [mh animals] NOT [mh humans] | **6790** |
| 11 | #9 NOT #10 | **74** |
