## Supplementary material for "Covid-19 in end-stage renal disease patients with renal replacement therapies: a systematic review and meta-analysis": Forest plots with subgroup analysis: ICU admission rate

### S3 Fig. Forest plots with subgroup analysis: ICU admission rate

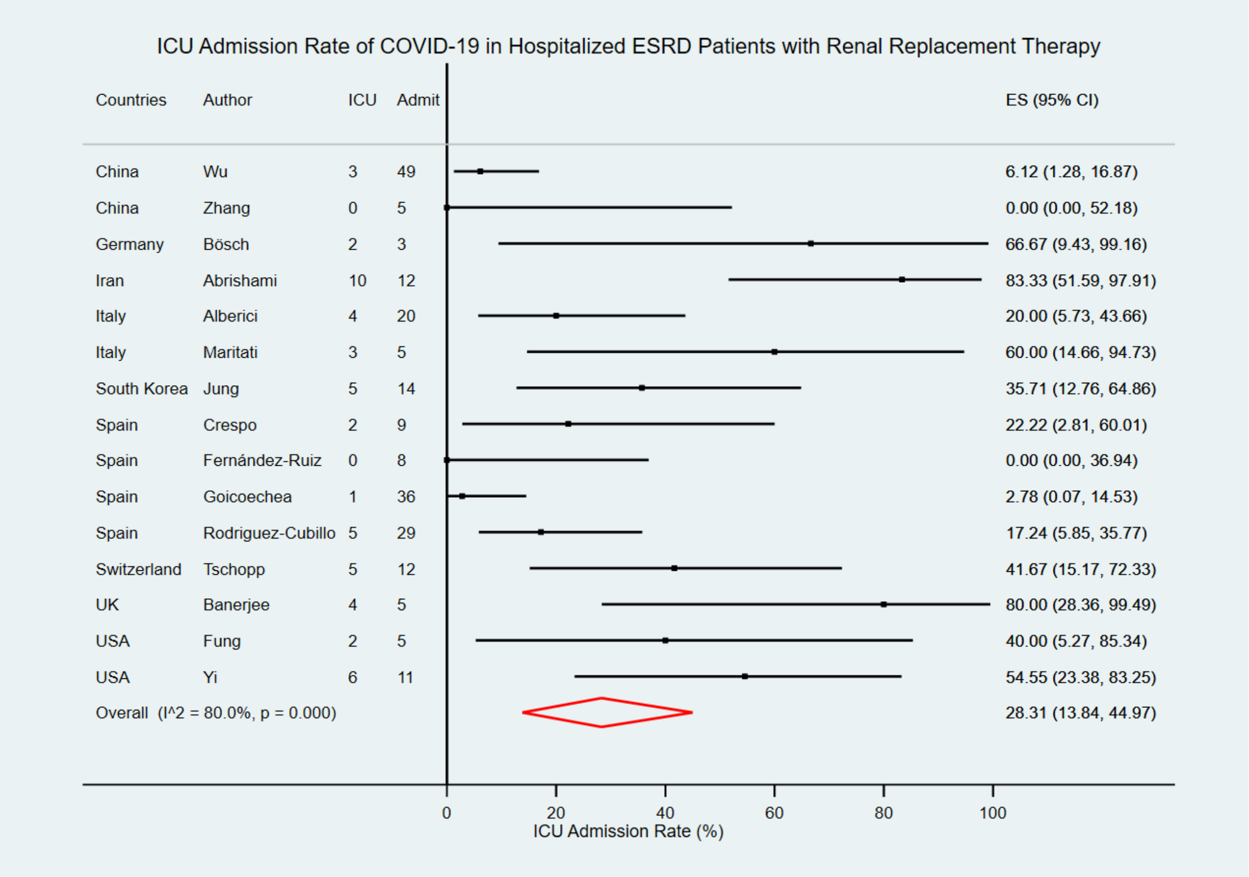


**S3A Fig. Forest plot: ICU admission rate of COVID-19 in hospitalized ESRD patients with RRT.** The figure summarizes the number of hospitalized COVID-19 cases with ICU admission in ESRD patients with RRT and the number of hospitalized COVID-19 cases in ESRD patients with RRT in 15 eligible studies. The forest plot represents the estimated ICU admission rate of COVID-19 in hospitalized ESRD patients with RRT for each study (black boxes), with 95% confidence intervals (95% CI; horizontal black lines). The overall estimated pooled ICU admission rate (red diamond) was 28.31% (95% CI = 13.84, 44.97%). The meta-analysis used a random-effects model with the exact method for confidence interval estimation. ES, effect size. $I^2$, test for heterogeneity.

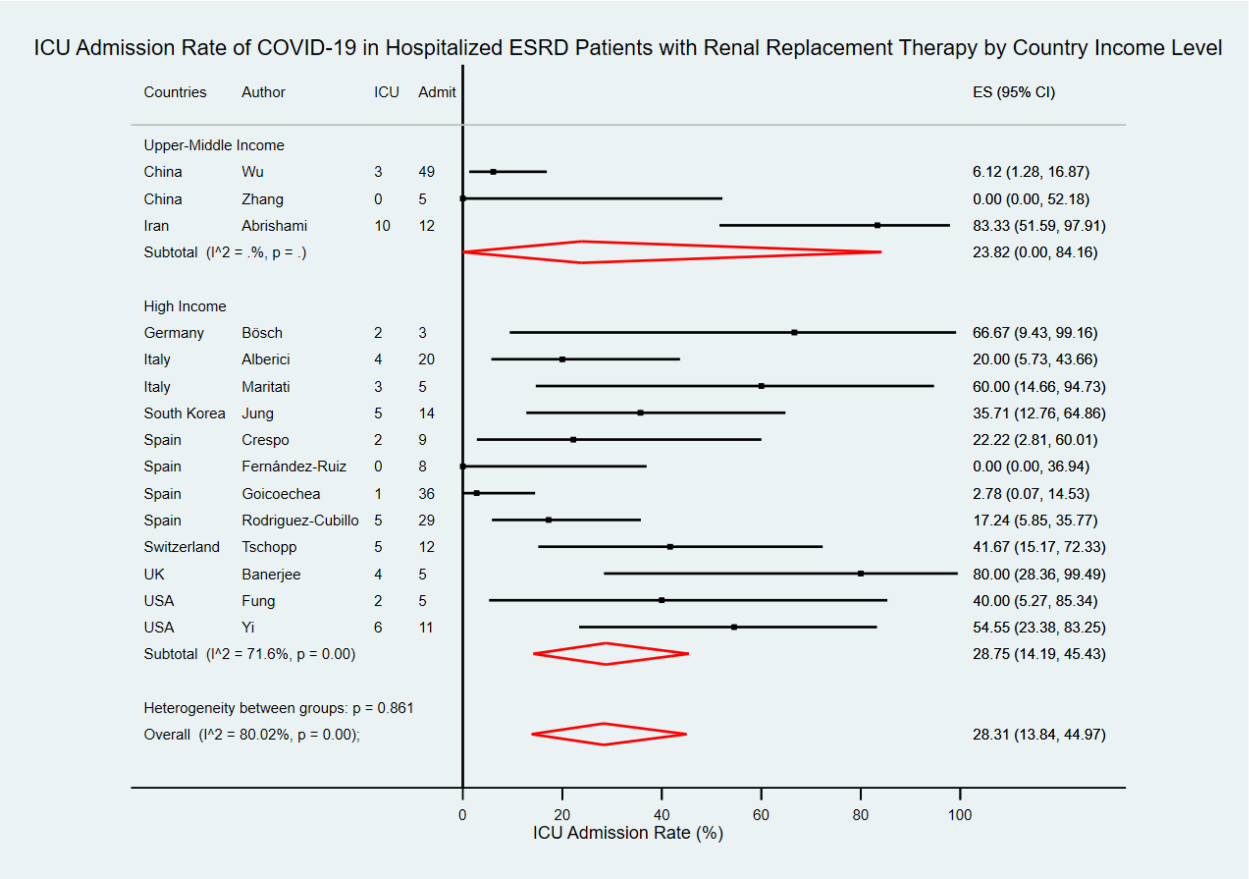


**S3B Fig. Forest plot: ICU admission rate of COVID-19 in hospitalized ESRD patients with RRT by country income level.** The figure summarizes the number of hospitalized COVID-19 cases with ICU admission in ESRD patients with RRT and the number of hospitalized COVID-19 cases in ESRD patients with RRT in 15 eligible studies with subgroup analysis by the World Bank country income level. The forest plot represents the estimated ICU admission rate of COVID-19 in hospitalized ESRD patients with RRT for each study (black boxes), with 95% confidence intervals (95% CI; horizontal black lines). The estimated pooled ICU admission rate for each subgroup was presented with a red diamond. The overall estimated pooled ICU admission rate (last red diamond) was 28.31% (95% CI = 13.84, 44.97%). The meta-analysis used a random-effects model with the exact method for confidence interval estimation. ES, effect size. $I^2$, test for heterogeneity.

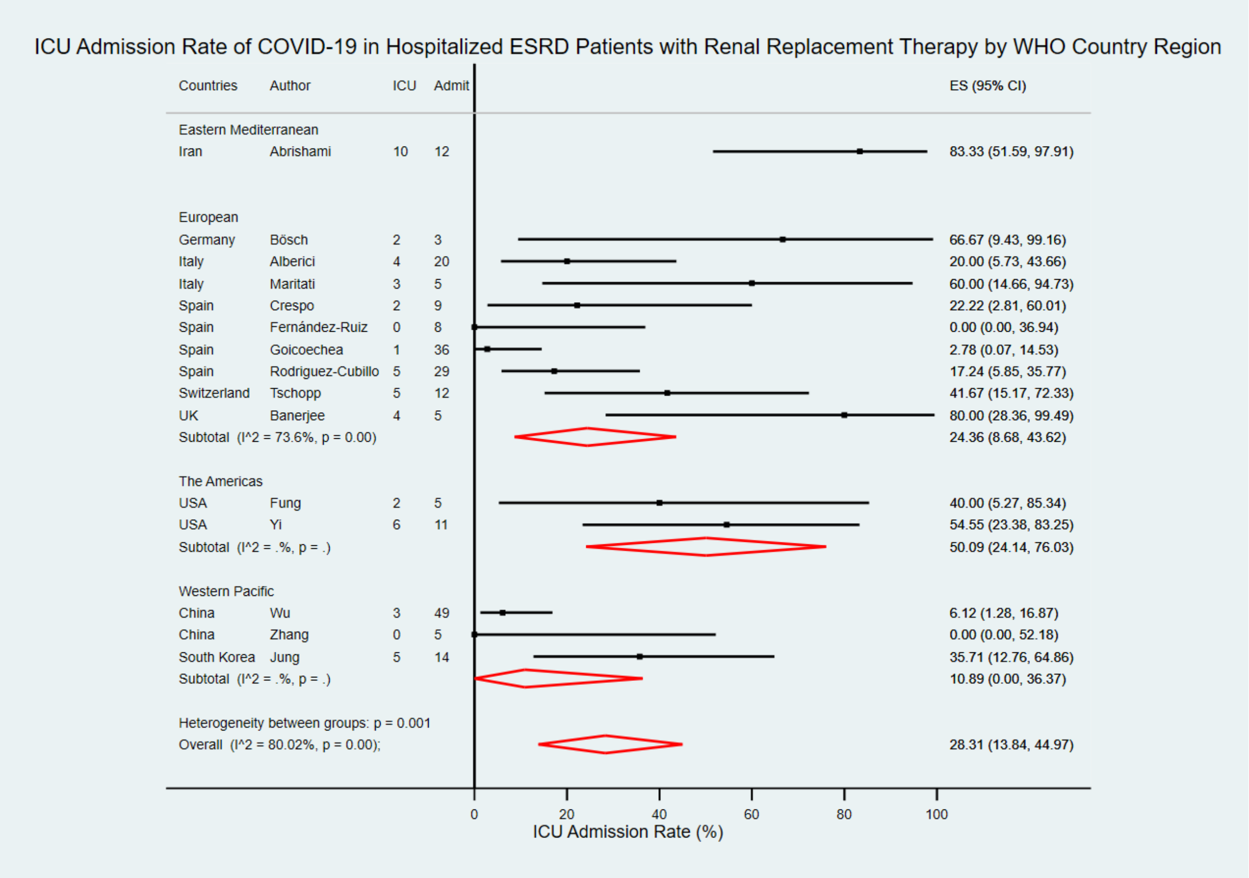


**S3C Fig. Forest plot: ICU admission rate of COVID-19 in hospitalized ESRD patients with RRT by WHO country region.** The figure summarizes the number of hospitalized COVID-19 cases with ICU admission in ESRD patients with RRT and the number of hospitalized COVID-19 cases in ESRD patients with RRT in 15 eligible studies with subgroup analysis by WHO country regions. The forest plot represents the estimated ICU admission rate of COVID-19 in hospitalized ESRD patients with RRT for each study (black boxes), with 95% confidence intervals (95% CI; horizontal black lines). The estimated pooled ICU admission rate for each subgroup was presented with a red diamond. The overall estimated pooled ICU admission rate (last red diamond) was 28.31% (95% CI = 13.84, 44.97%). The meta-analysis used a random-effects model with the exact method for confidence interval estimation. ES, effect size. $I^2$, test for heterogeneity.

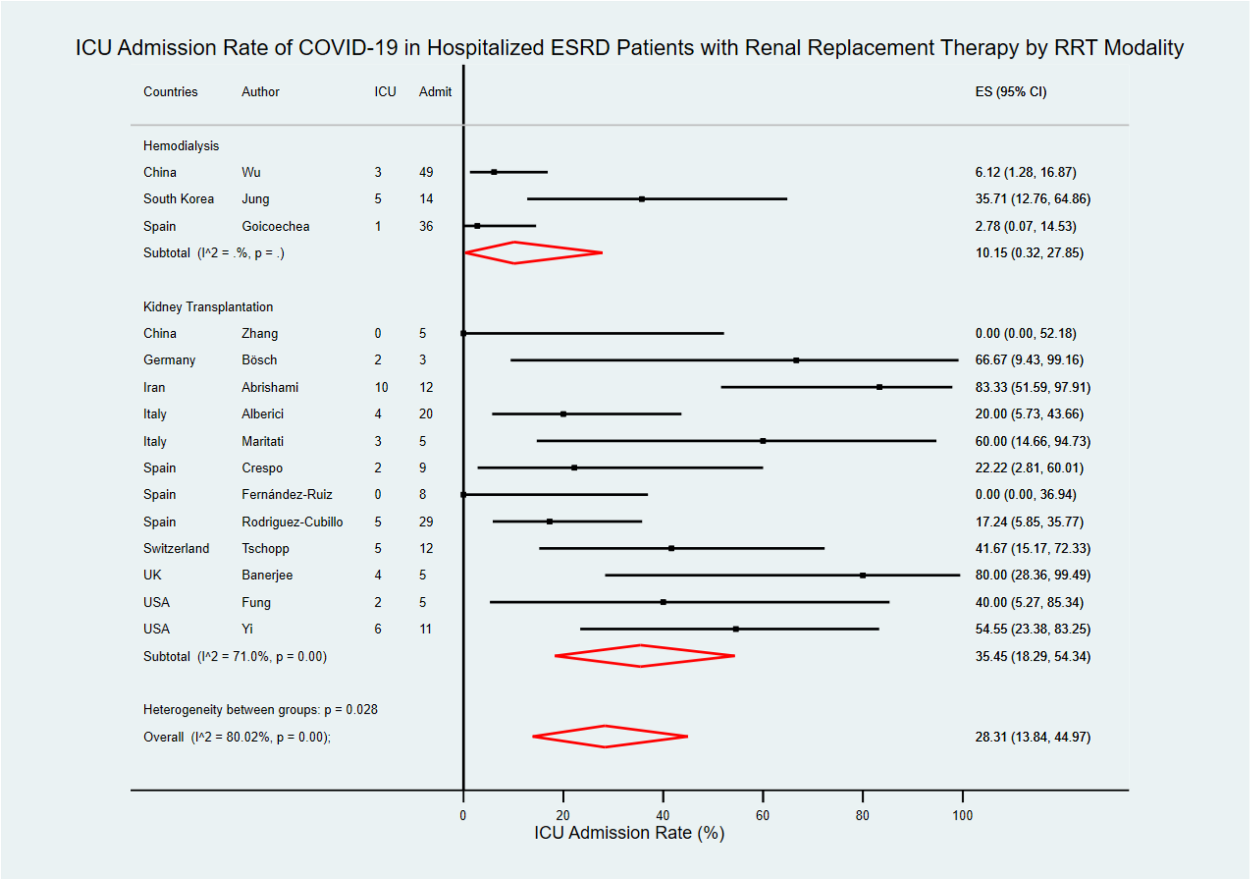


**S3D Fig. Forest plot: ICU admission rate of COVID-19 in hospitalized ESRD patients with RRT by RRT modality.** The figure summarizes the number of hospitalized COVID-19 cases with ICU admission in ESRD patients with RRT and the number of hospitalized COVID-19 cases in ESRD patients with RRT in 15 eligible studies with subgroup analysis by types of RRT modality. The forest plot represents the estimated ICU admission rate of COVID-19 in hospitalized ESRD patients with RRT for each study (black boxes), with 95% confidence intervals (95% CI; horizontal black lines). The estimated pooled ICU admission rate for each subgroup was presented with a red diamond. The overall estimated pooled ICU admission rate (last red diamond) was 28.31% (95% CI = 13.84, 44.97%). The meta-analysis used a random-effects model with the exact method for confidence interval estimation. ES, effect size. $I^2$, test for heterogeneity.
